# Fragment end motif analysis to distinguish pathogens from contaminants in enriched plasma microbial DNA

**DOI:** 10.1101/2025.11.06.25339688

**Authors:** Haikun Zhang, Eddie G. Dominguez, Mary Junak, Muhammed Murtaza, Caitlin S. Pepperell, Mehreen T. Kisat

## Abstract

**Introduction:** Despite its promise, accuracy of microbial cell-free DNA (mDNA) in plasma as a diagnostic tool is hindered by its low abundance and process contaminants. We have previously shown that combining size selection with single-stranded DNA (ssDNA) library preparation increased mDNA yield by 200-fold but also decreased sensitivity for pathogen detection due to higher background noise. A recent study showed that pathogen-derived DNA was enriched for CC dinucleotide at 5’ ends compared to contaminants. Since ssDNA libraries preserve sequence motifs at both ends (5’ and 3’), we hypothesized that analysis of nucleotide motifs at microbial fragment ends in size-selected ssDNA libraries could help differentiate pathogen DNA from background noise.

**Methods:** We performed deep sequencing on size-selected ssDNA libraries (<110 bp) generated from longitudinal plasma samples of 11 critically-ill patients (5 with culture-proven infections, 20 samples; 6 without infections, 18 samples) and 6 no-template controls (NTCs). For each 2-mer and 1-mer motif, we calculated the ratio between its frequency observed at 5’ and 3’ fragment ends in sequencing data and its expected frequency in the corresponding reference genome (O/E ratio). We compared enrichment of motifs in pathogen DNA and contaminant DNA fragments.

**Results:** Pathogen-derived mDNA fragments were more biased in O/E end motif ratios compared to contaminants across all 3 groups (NTCs, no-infections and culture-proven infections), at both 5’ and 3’ fragment ends. Notably, the GG dinucleotide was enriched at the 3’ end in pathogens compared to contaminants (P < 0.0001). Combining O/E ratios for C and G nucleotides at the 3’ end achieved areas under the receiver operating characteristic curve of >0.98 for distinguishing common contaminants from culture-proven pathogens.

**Conclusions:** Pathogen-derived mDNA in size-selected ssDNA libraries is biased at 5’ and 3’ fragment end compared to contaminants. Incorporating microbial fragment end motif analysis can enhance signal-to-noise ratio and improve pathogen detection and identification in plasma metagenomic sequencing.

## Introduction

Sepsis, which is characterized by a dysregulated host inflammatory response to infection is a leading cause of death in critically ill patients^1–3^. Sepsis develops in 20-25% of trauma patients admitted to the Intensive Care Unit (ICU) and is associated with a mortality rate of 20%^1,2^. Differentiating sepsis from sterile systemic inflammatory response remains challenging in trauma patients,^4,5^ due to the overlap in clinical signs and symptoms between injury associated inflammation and sepsis^6^. To err on the side of caution, we start patients on broad spectrum antimicrobial treatment while awaiting microbiology culture results^7^. This practice leads to antimicrobial resistance and opportunistic infections such as *C. difficile* colitis^8^. We continue empiric treatment for 3-5 days due to the long turnaround times for final microbiology cultures. Even then, cultures fail to identify causative pathogens in more than 30% of patients with sepsis^9,10^. Thus, there is an unmet need for rapid and accurate identification of sepsis and its causative pathogens.

Cell-free DNA (cfDNA) in the bloodstream comes from host cells as well as from commensal or invasive microbes that release microbial DNA (mDNA) into circulation^11,12^. The ability to use a test based on the presence of mDNA fragments in plasma for infectious disease diagnostics is promising as it can be less invasive and faster compared to conventional microbiology methods (24-48 hours versus 3-5 days respectively)^12–14^. We and others have demonstrated that mDNA can be detected in patients with ongoing infections using plasma metagenomic sequencing^15^. This approach can be used for sepsis diagnosis and pathogen identification^13,16^. However, there are several technical limitations before an mDNA test can be used routinely at the bedside in the ICU. One such limitation is the very small ratio of microbial to human fragments in plasma DNA (1:1000), making it challenging and expensive to obtain sufficient microbial DNA for sequencing-based analysis.

To address this limitation, we recently developed an approach combining single-stranded DNA (ssDNA) library preparation (captures smaller and more fragmented DNA) with fragment size selection (< 110 bp, depletes human cfDNA). This method enriches mDNA yield (signal) by about 200-fold on average, improving recovery and sequencing resolution for microbial DNA in plasma, while lowering sequencing costs^17^. However, even as we observed an increase in microbial cfDNA yield, the sensitivity of pathogen detection decreased due to higher background contribution from equally enriched contaminant and commensal microbial DNA (noise). Thus, to utilize enrichment to improve mDNA analysis as a reliable and cost-effective clinical biomarker for diagnosis and management of sepsis, improvement in signal-to-noise ratio (SNR) is necessary.

There are several strategies that have been used to improve SNR in plasma metagenomic sequencing, with most studies focused on tracking and removing background noise from contaminant genera (such as using no template controls), or comparing abundance of genera between samples, such that an outlier microbe is considered a true-positive pathogen.^15,18^. While these approaches reduce background noise, they lead to false-negative results for genera that are common as contaminants, but can also cause infections. To overcome this challenge, a recent study has shown that pathogen-derived microbial cfDNA is enriched for “CC” dinucleotides at fragment ends compared with contaminant-derived DNA, suggesting in vivo enzymatic digestion^19^. This approach can potentially distinguish true pathogens from background contaminants. However, this study was limited primarily to patients with bacteremia and a small number of pathogens, and a relatively high abundance of microbial DNA in plasma. In addition, the study used double-stranded DNA (dsDNA) library preparation which does not capture single stranded microbial DNA fragments or fragments with nicks or overhangs, and only preserves sequence motifs at 5’ ends since it requires end repair (filling of 5’ overhangs and excision of 3’ overhangs)^20^. In contrast, ssDNA library preparation allows sequencing of the diverse pool of mDNA fragments, and preserves both 5’ and 3’ ends^20,21^, enabling study of DNA fragment end motif patterns previously not described in microbial cfDNA metagenomic sequencing. Here, we evaluate end motif signatures from enriched single-stranded cfDNA libraries for microbial DNA analysis. We hypothesize that analysis of nucleotide motifs at 5’ and 3’ microbial fragment ends in size-selected ssDNA libraries will allow differentiation of pathogen DNA from background noise.

## Methods

### Study cohort and sample collection

Patients were enrolled in an on-going study (January 2022-March 2025) recruiting trauma patients admitted to the intensive care unit (ICU) from the Emergency Department at University of Wisconsin Hospitals and Clinics. Longitudinal blood samples were collected for the first 10 days after ICU admission. To evaluate pathogen detection, we included 5 patients with sepsis and 6 patients without infection (confirmed by expert consensus of a multidisciplinary board-certified team). From patients with sepsis, we specifically selected 20 samples for deeper sequencing in the current study because they were collected within 1 day of a positive microbial culture or were previously shown to have high pathogen DNA burden. The study was approved by the institutional review board of the University of Wisconsin-Madison (IRB protocol number 2021-0484). Written informed consent was obtained from participants or their legal designees.

### Blood processing and cell-free DNA extraction

Blood samples were collected and processed using laboratory protocols optimized for cell-free DNA analysis^22^. At each time point, up to two 10 ml Streck Cell-Free DNA blood tubes were obtained. Blood samples were processed within 24 hours of collection with two rounds of centrifugation to isolate plasma (820g for 10 min; 20,000g for 10 min). Plasma samples were stored at -80°C in 1 ml aliquots until further use. DNA was extracted using the ThermoFisher MagMAX Cell-Free DNA extraction kit according to the manufacturer’s protocol.

### Single-stranded library preparation, size selection and sequencing

Sequencing libraries were prepared using a single-stranded (SRSLY NGS Library Prep Kit; Claret Bioscience) DNA kit followed by size selection using automated agarose gel electrophoresis (Blue Pippin, Sage Science) to exclude DNA fragments longer than 110 bp (Figure 1) as previously described^17^. Additionally, we performed 4 cycles of PCR amplification using a commercially available next-generation sequencing master mix (Invitrogen^TM^ Collibri^TM^ Library Amplification Master Mix with Primer Mix) on the library pool prior to size selection.

**Figure 1.**
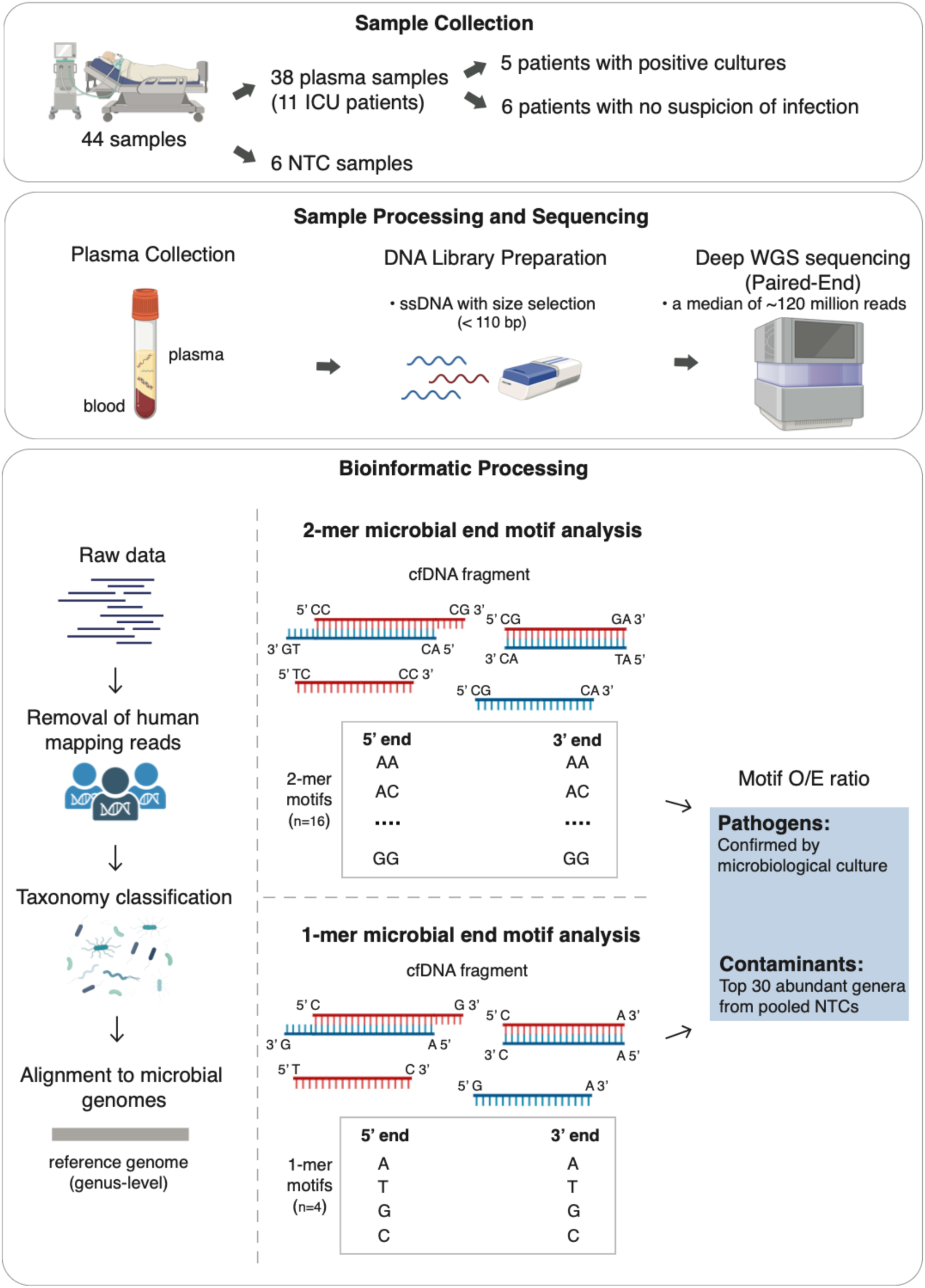
Overview of patient cohort, sample processing, and computational workflow. A total of 38 plasma samples were collected from 11 ICU patients, including 20 samples from 5 patients with culture-proven infections and 18 samples from 6 patients without infection. 6 no-template control (NTC) samples were processed through sampling processing to evaluate potential contamination. Plasma samples were processed using magnetic bead-based extraction methods, followed by single-stranded DNA (ssDNA) library preparation and size-selection to exclude fragments > 110 bp. After paired-end deep sequencing, human-mapped reads were removed, and the remaining unmapped reads were taxonomically classified using Kraken2 and Bracken. Microbial fractions were quantified at both total and pathogen-specific levels for downstream analysis. For microbial end motif analysis, reads classified to each genus were re-aligned to their corresponding reference genomes using BWA. Dinucleotide (2-mer) and mononucleotide (1-mer) microbial end motif analysis was used to differentiate pathogens from contaminants.

To assess for contaminants, 6 no-template control (NTC) samples were included for the entire processing workflow, including cfDNA extraction, library preparation and fragment size selection. All DNA libraries after size selection were sequenced on an Illumina NextSeq 2000 platform with paired-end reads (2 x 50 bp) using the P4 flowcell.

### Sequencing data processing and alignment

After base calling, paired-end fastq files were trimmed using fastp (v0.20.1)^23^ to remove adaptor sequences and low-quality bases with default parameters. Filtered paired-end reads were aligned to the human reference genome hgT2T (v2.0) using BWA (v0.7.17)^24^. Human reads were removed with SAMtools (v1.15.1)^25^. Read pairs with at least one read that unmapped to the human genome were extracted using SAMtools with the parameters “--rf 12 -bhF 3840”. These unmapped reads were used for microbial taxonomic classification.

### Microbial taxonomic classification and pathogen-specific DNA fraction calculation

Taxonomic classification was performed using Kraken2 (v2.1.2)^26^, a k-mer based taxonomic classifier, with the prebuilt Refseq Kraken2 database. Reads classified to specific genus by Kraken2 were extracted for downstream end motif analysis. To estimate microbial fraction, genus and phylum level read counts from Kraken2 were further refined using Bracken (v2.7)^27^ with the parameters “-r 50 -t 2”. Microbial fragments were defined as fragments assigned to any microbial phylum over filtered sequencing fragments. For each microbe confirmed by microbiological culture, fragments classified to the genus of the identified pathogen were counted. The pathogen-specific DNA fraction was calculated as the ratio of fragments assigned to the pathogen’s genus to the total microbial fragments. Additionally, reads per million (RPM) were defined as the ratio of fragments assigned to the genus over all the total cfDNA fragments (∼M reads).

### Microbial end motif analysis

Microbial end motif analysis was performed at the genus level using reads classified to the specific genus by Kraken2. The microbial reads classified to each genus were re-aligned to their corresponding reference genomes using BWA at genus level. Genomic positions and aligned sequences were obtained using Bedtools (v2.30.0)^28^.

To characterize end motifs, nucleotide sequences (1-mer and 2-mer) from both 5’ (R1) and 3’ (R2) fragment ends were analyzed. The frequency of each end motif was calculated as the counts of that motif divided by the total number of all end motifs for the specific microbial genus. As microbial end motif frequencies are influenced by reference genome sequence composition^19^, we applied sequence-based normalization by computing the ratio of observed to expected end motif frequency (O/E ratio) within each genus:

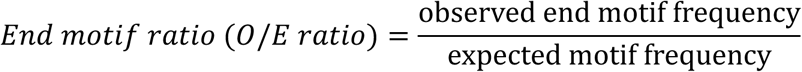

The observed end motif frequency refers to the mDNA end motif frequency of specific genus derived from cfDNA sequencing data, while the expected motif frequency was defined as the motif frequency of the specific microbial reference genome, which was calculated in silico by scanning reference genomes (both strands) using 1-mer or 2-mer sliding windows. An O/E ratio near 1 indicates unbiased use of the end motif during DNA fragmentation, while higher or lower O/E ratios suggest enrichment or depletion of specific end motifs, respectively. For genera involving multiple reference genomes, expected motif frequencies were estimated using a weighted average across genomes, with weights based on the proportion of reads mapped to each reference genome within the specific genus:

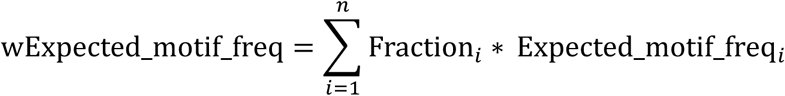

where wExpected_motif_freq represents the weighted reference motif frequency for a specific genus. Fraction_i_ is the proportion of reads mapped to the reference genome i within that genus. Expected_motif_freq_i_ refers to the motif frequency of reference genome i involving in the specific genus.

The 2-mer CG O/E ratio was a combination of CC, CG, GC and GG end motif O/E ratio, which was defined as:

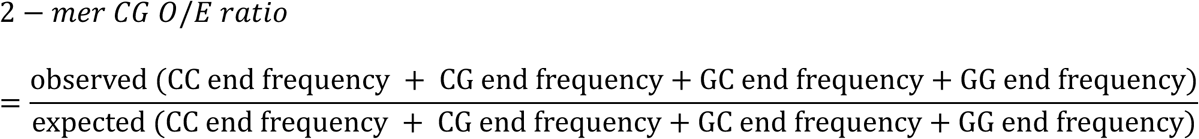

And the 1-mer CG O/E ratio as:

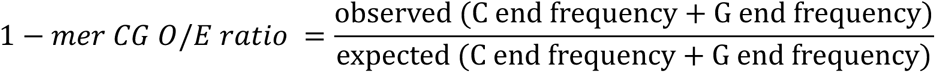

The fraction of pathogen-specific mDNA with a specific end motif was calculated as the ratio of fragments from the pathogen genus with that motif to the total microbial reads.

### Statistical analysis

The Mann-Whitney U test was used to compare end motif O/E ratios between pathogens from sepsis samples and top 30 contaminating genera from NTCs. End motif variability within each genus was quantified using coefficient of variation (CV) of all 16 2-mer O/E ratios. The AUCs measuring the discrimination between pathogens and contaminants from different groups (NTCs, non-Infection and sepsis) were performed using a logistic regression-based prediction model. Analysis of ROC curves was constructed using R package PredictABEL. All data analyses were performed using Perl and R.

## Results

### Study participants and samples information

We analyzed 38 longitudinal plasma samples collected from 11 critically ill trauma patients admitted to the intensive care unit (ICU). Among them, 5 patients had culture-confirmed infections (sepsis group; 20 samples) and 6 patients were confirmed without infection (non-infection group; 18 samples; confirmed by expert consensus of a multidisciplinary team of clinical experts) (Figure 1). Across the 5 sepsis patients, a total of 33 microbiological cultures were obtained (blood, urine, and bronchoalveolar lavage/tracheal aspirate/sputum), of which 8 were positive (*Staphylococcus aureus* = 5, *Streptococcus pneumoniae* = 1, *Gardnerella vaginalis* = 1, *Klebsiella (Enterobacter) aerogenes* = 1, *Haemophilus influenzae* = 2, *Serratia marcescens/ureilytica* = 1) (Supplemental Table 1). Of the 20 samples from sepsis patients selected for this analysis, 17 were coincident with positive cultures (obtained day of positive culture or within 1 calendar day), and 3 were collected at time points previously shown to have a high burden of pathogen-specific DNA based on shallow sequencing^17^. Additionally, 6 no-template control (NTC) samples were processed through the entire process from cfDNA extraction and library preparation to assess potential contamination.

### Size-selected single-stranded library preparation for mDNA enrichment

We previously developed a size-based enrichment strategy for mDNA, demonstrating that combining single-stranded DNA (ssDNA) library preparation with fragment size selection (< 110 bp) significantly increased the mDNA fraction by an average of 200-fold using shallow metagenomic sequencing^17^. In this study, we performed deep metagenomic sequencing on size-selected ssDNA libraries for plasma samples, generating a median sequencing depth of 122 million reads per library (interquartile range [IQR]: 64-295 million) after quality control. Following quality control, filtered sequencing reads were aligned to the human reference genome hgT2T, and unaligned reads were used for taxonomic classification. Using Kraken2 and Bracken to perform taxonomic classification, we were able to classify a mean proportion of 0.0057 (SD = 0.0054) of all sequenced fragments from plasma samples and 0.127 (SD = 0.014) from NTC samples as microbial at the phylum level (Supplemental Figure 1). No significant difference in total microbial abundance was observed between sepsis and non-infection samples (Mann-Whitney U test, p = 0.196; Supplemental Figure 1).

### Pathogen detection based on microbial abundance in size-selected ssDNA libraries

A total of 17 samples from 5 sepsis patients were coincident with 8 culture-proven infections (6 unique bacterial species; *Staphylococcus aureus*, *Streptococcus pneumoniae*, *Gardnerella vaginalis*, *Klebsiella aerogenes*, *Haemophilus influenzae*, *Serratia marcescens/ureilytica*). Among these, *Staphylococcus* was the most prevalent pathogen genus, accounting for 38.5% (10 of 26). We observed presence of genera for some of the cultured pathogens in NTC and non-infection samples (e.g., *Staphylococcus*; Figure 2). Pathogen-specific abundance was determined at the genus level as the fraction of genus-specific reads over the total microbial reads. Using the 18 non-infection samples as background, 10 of 26 (38.5%) pathogens were detected significantly above background. We observed that due to the overlap in abundance of pathogen genera in patients with infection, patients without infection and in NTCs, it is difficult to distinguish infection-causing pathogens from contaminants and commensals using microbial abundance alone.

**Figure 2.**
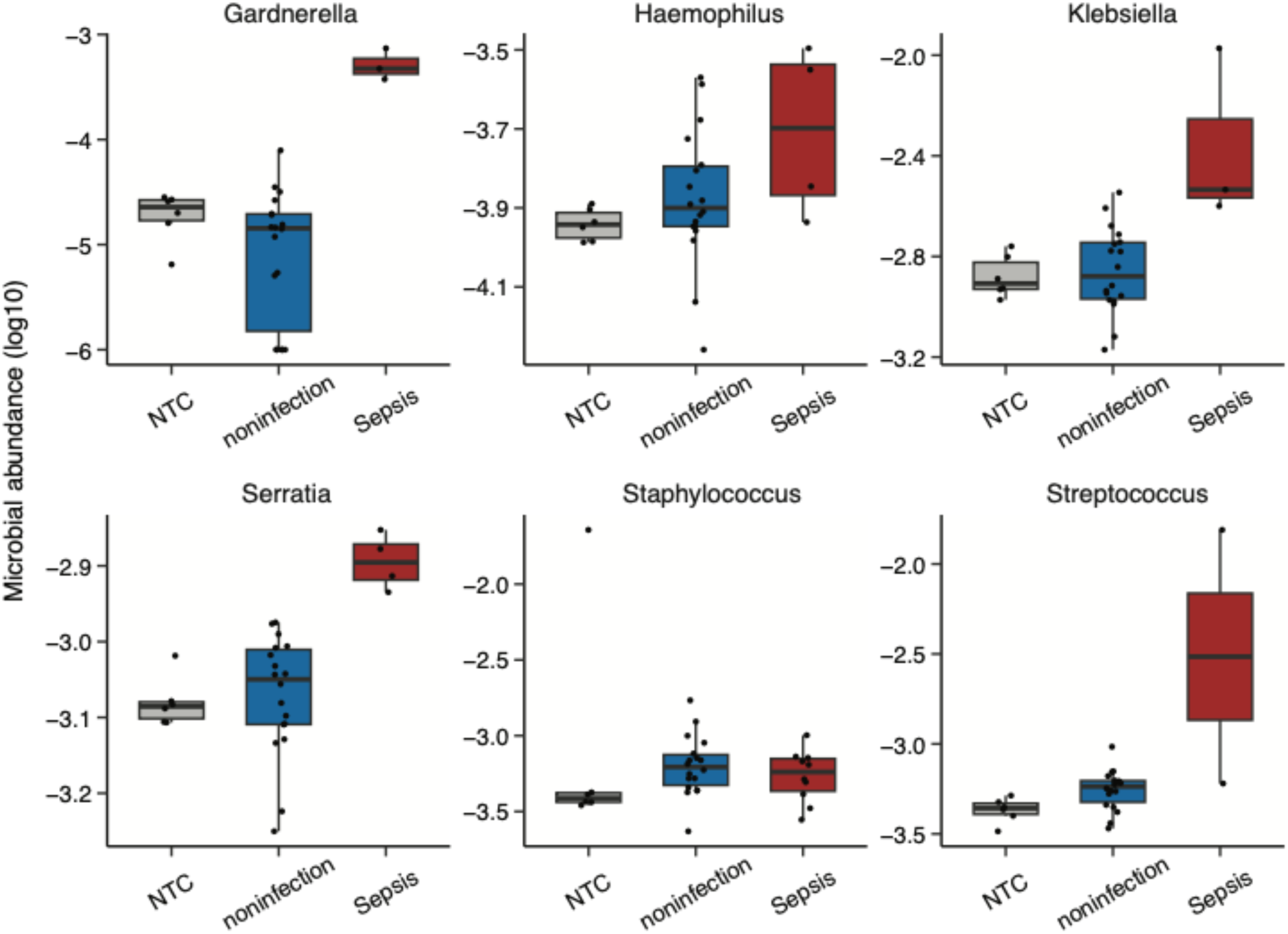
Pathogen-specific microbial abundance (genus-level) across sepsis, non-infection and NTC samples. Boxplots show the relative abundance of pathogen-associated genera detected in plasma cfDNA across three groups. Pathogen-specific microbial abundance was calculated as the ratio of fragments assigned to each pathogen genus to the total microbial fragments in the sample. Pathogens in sepsis samples correspond to those confirmed by microbiology culture at coincident time points.

### Plasma microbial end motif analysis

To characterize the end motif profiles of contaminating microbial fragments, we pooled six NTC samples and identified the 30 most abundant microbial genera. The abundances of contaminating microbial genera in size-selected ssDNA was substantially higher than that typically observed in double-stranded libraries^17^ (i.e., *Pseudomonas* showed a median of 195,505 sequences). The microbial fragments within the top 30 genera were considered contaminating microbial DNA and used for downstream end motif analysis. The O/E ratios for 2-mer end motifs across contaminating genera were close to 1 (Figure 3), with minimal variability at both 5’ (median and IQR of CV, 0.143 [0.128-0.173]) and 3’ ends (median and IQR of CV, 0.2 [0.184-0.212]), suggesting a random fragmentation pattern of contaminating microbial DNA in NTCs.

**Figure 3.**
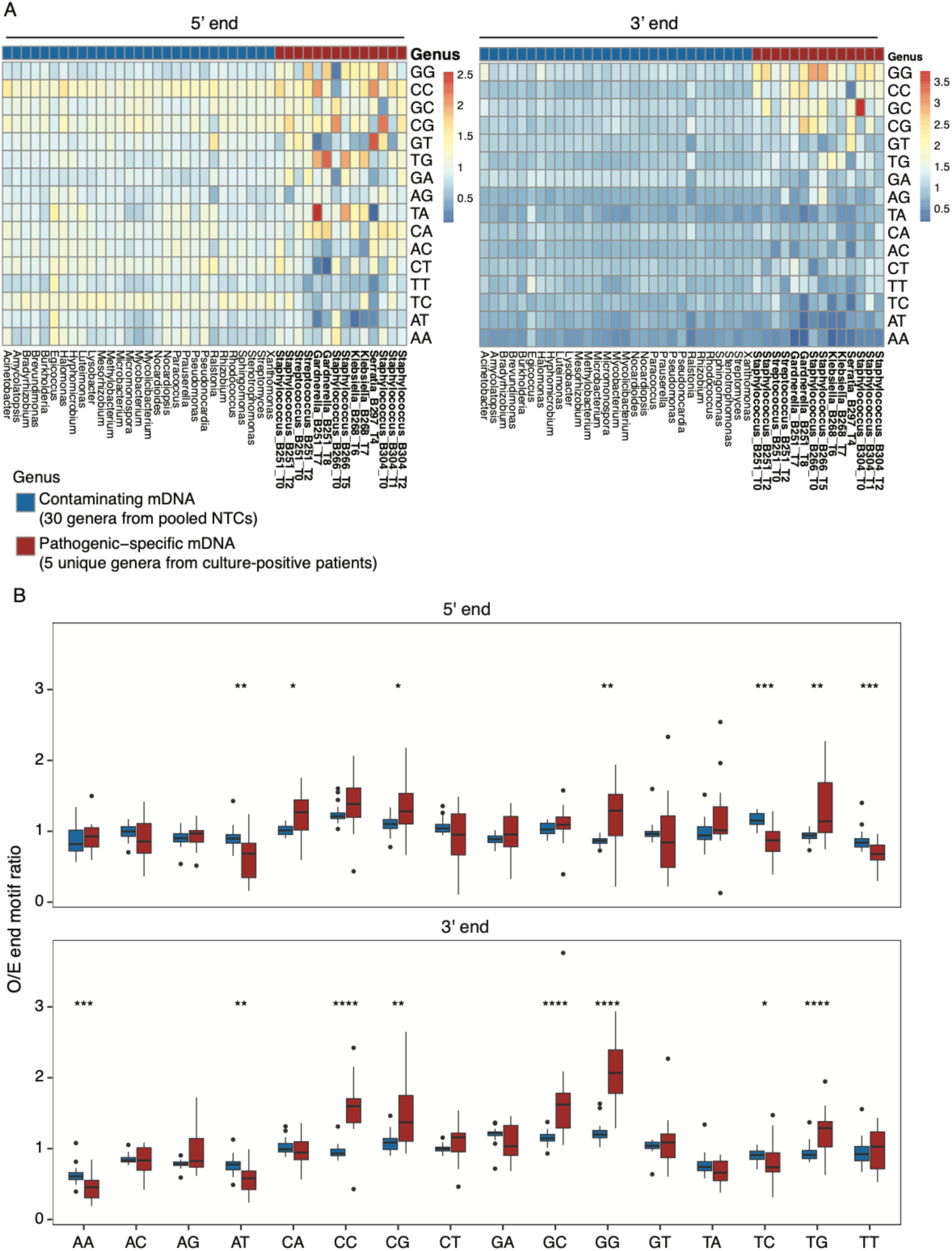
Dinucleotide microbial end motif patterns at the 5’ and 3’ fragment ends. (A) Heatmap shows the O/E ratios of 2-mer end motifs for pathogenic genera (five unique genera from culture-positive patients) and contaminating genera (top 30 abundant genera from pooled NTCs). Motif patterns are shown separately for the 5’ and 3’ fragment ends. (B) Boxplots compare the O/E ratios of all 16 2-mer end motifs between pathogen-derived and contaminant-derived mDNA at both fragment ends (*p <0.05; **p < 0.01; ***p < 0.001; **p < 0.0001).

### Dinucleotide microbial fragment profiles at 5’ and 3’ ends distinguish culture-proven pathogens from contaminants

We further analyzed 2-mer end motif profiles at both the 5’ and 3’ fragment ends to compare pathogen-derived microbial cfDNA and contaminating microbial DNA. The O/E ratios of all 2-mer end motifs were calculated for the top 30 microbial genera identified in pooled NTCs (contaminants) and for pathogens confirmed by microbiological culture in sepsis samples. To ensure reliable analysis, only genera with more than 200 fragment counts (14 pathogenic genera) were included, as described in an earlier study^17^.

Distinct end motif patterns were observed between pathogen-derived cfDNA from sepsis samples and contaminating microbes in NTCs at both the 5’ and 3’ end (Figure 3A). While contaminants exhibited motif patterns indicative of random fragmentation, pathogen-derived mDNA showed enrichment for specific motifs at both termini. Notably, the difference in end motif O/E ratios between pathogens and contaminants was greater at the 3’ end (median and IQR of CV, 0.468 [0.403-0.553] vs. 0.2 [0.184-0.212]; p=1.02 x10^-10^, Mann-Whitney U test) than at the 5’ end (median and IQR of CV, 0.415 [0.264-0.494] vs. 0.143 [0.128-0.173]; p=1.71 x10^-9^, Mann-Whitney U test;) (Figure 3A, Figure 4A). Differential end motifs (DEMs) were then identified between pathogens and contaminants for both the 5’ and 3’ ends (Figure 3B, Supplementary Figure 2). At the 5’ end, GG and TG motifs were significantly enriched, whereas AT, TC and TT were depleted in pathogens. At the 3’ end, GG, CC, GC, CG and TG motifs were significantly enriched, while AA and AT were depleted relative to contaminants.

**Figure 4.**
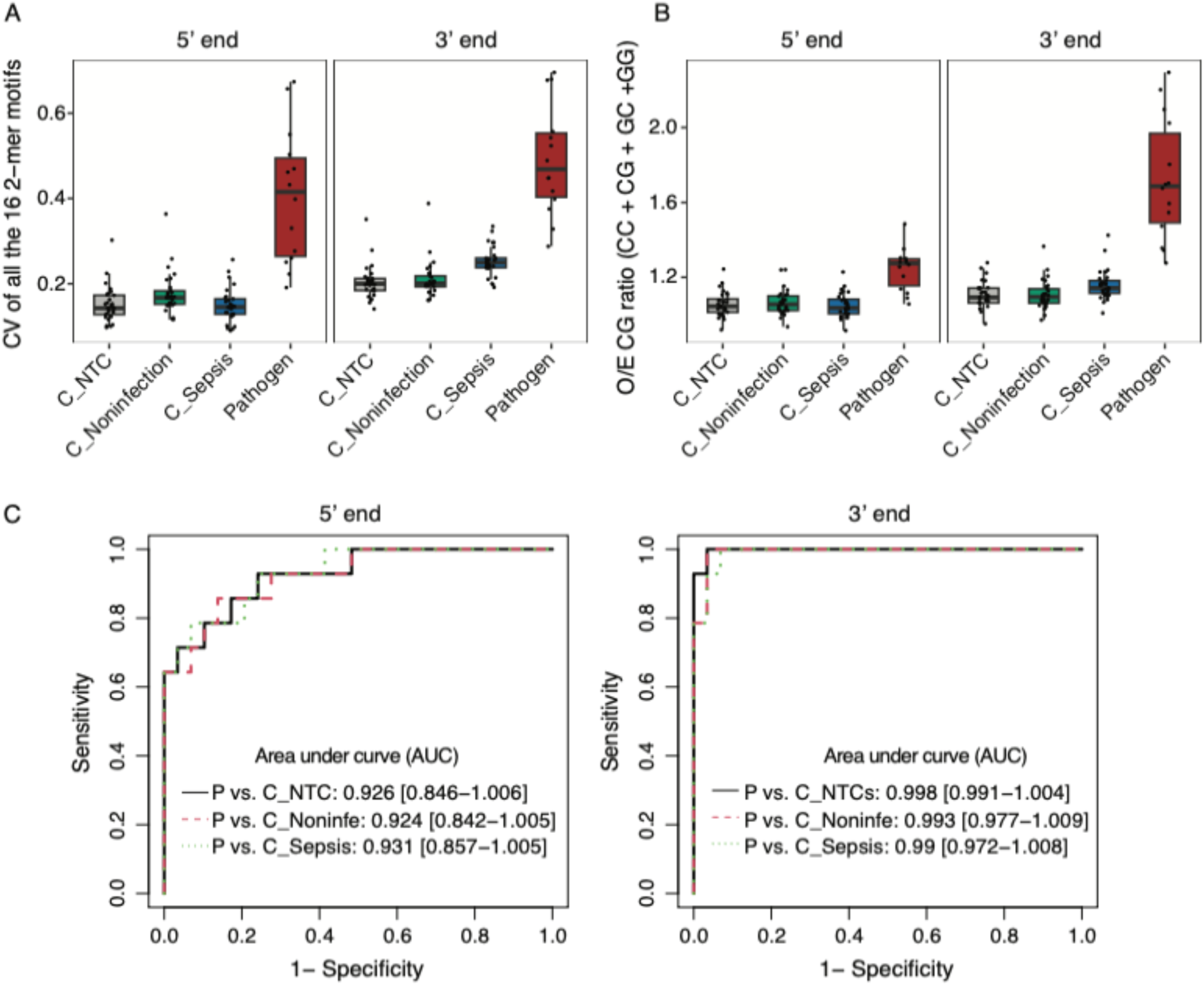
Comparative analysis of 2-mer microbial end motif patterns in pathogen- and contaminant-derived mDNA. (A) Coefficient of variation (CV) of 2-mer end motifs in culture-proven pathogens from sepsis samples and contaminants (top 30 genera identified from pooled NTCs) across pooled NTC, non-infection and sepsis groups. (B) the O/E CG ratios (combining GG, GC, CG and CC motifs) compared between pathogens and contaminants across pooled NTC, non-infection and pooled groups. (C) Receiver operating characteristic (ROC) curves showing the performance of the combined CG ratio in differentiating pathogens from contaminants across different groups. Contaminants were defined as the 30 most abundant genera from pooled NTCs. C_NTC, contaminants from pooled NTC samples; C_Noninfection, contaminants from pooled non-infection samples; C_Sepsis, contaminants from pooled sepsis samples.

We next compared the end motif profiles of contaminating microbial DNA detected in plasma samples with culture-proven pathogens from sepsis samples. Since the top 30 microbial genera in NTCs were considered contaminants, the end motif profiles of these contaminating genera were further evaluated in pooled non-infection and sepsis samples. The median CV of all 16 motif O/E ratios within contaminating genera across pooled NTC, non-infection and sepsis samples were 0.143, 0.167, 0.145 for the 5’ end, and 0.2, 0.2, 0.25 for the 3’ end, respectively. In contrast, pathogen-derived mDNA showed markedly higher motif variability (median CV = 0.415 for 5’ end and 0.468 for 3’ end), significantly higher than that of contaminants (Figure 4A). These findings suggested that ssDNA fragment end patterns after size selection were different between pathogens and contaminants, particularly at the 3’ end.

Next, we combined the O/E ratios of the most overrepresented 2-mer motifs (GG, GC, CG and CC) into a CG ratio (Methods). The CG ratio was consistent across contaminants in NTC, non-infection and sepsis samples (Figure 4B, Supplemental Figure 3). Notably, the difference in CG ratio between pathogens and contaminants was larger at the 3’ end than at 5’ end. Receiver operating characteristic (ROC) analysis demonstrated that the CG ratio achieved an area under the ROC curve (AUC) of 0.92 at 5’ end and 0.99 at 3’ end in distinguishing culture-proven pathogens from contaminants across all sample groups (Figure 4C). Overall, these findings suggested that microbial end motif patterns, particularly at 3’ termini, may help discriminate true pathogens from potential contaminants introduced during sample processing.

### 3’-end mononucleotide signatures enable reliable differentiation of pathogens from contaminants

Given the low abundance of pathogen-specific mDNA in certain samples, we extended the end motif analysis to 1-mer resolution. The 1-mer based approach requires fewer fragments for reliable inference, thereby reducing overall sequencing depth while increasing the number of evaluable samples in subsequent comparative analyses. We first evaluated the performance of the 1-mer approach using the same samples in the 2-mer analysis to differentiate infection-causing pathogens from contaminants. Consistent with the 2-mer results, distinct end motif patterns were observed between pathogens from sepsis samples and contaminants in NTCs at both 5’ and 3’ end (Figure 5A). Pathogen-derived cfDNA showed significant enrichment of C and G termini at both ends, whereas A end was significantly depleted at the 3’ end (Figure 5B). Further analysis of contaminating microbial DNA in pooled non-infection and sepsis samples revealed lower variability in 1-mer end motifs compared with pathogen-derived mDNA from sepsis samples (Figure 5C).

**Figure 5.**
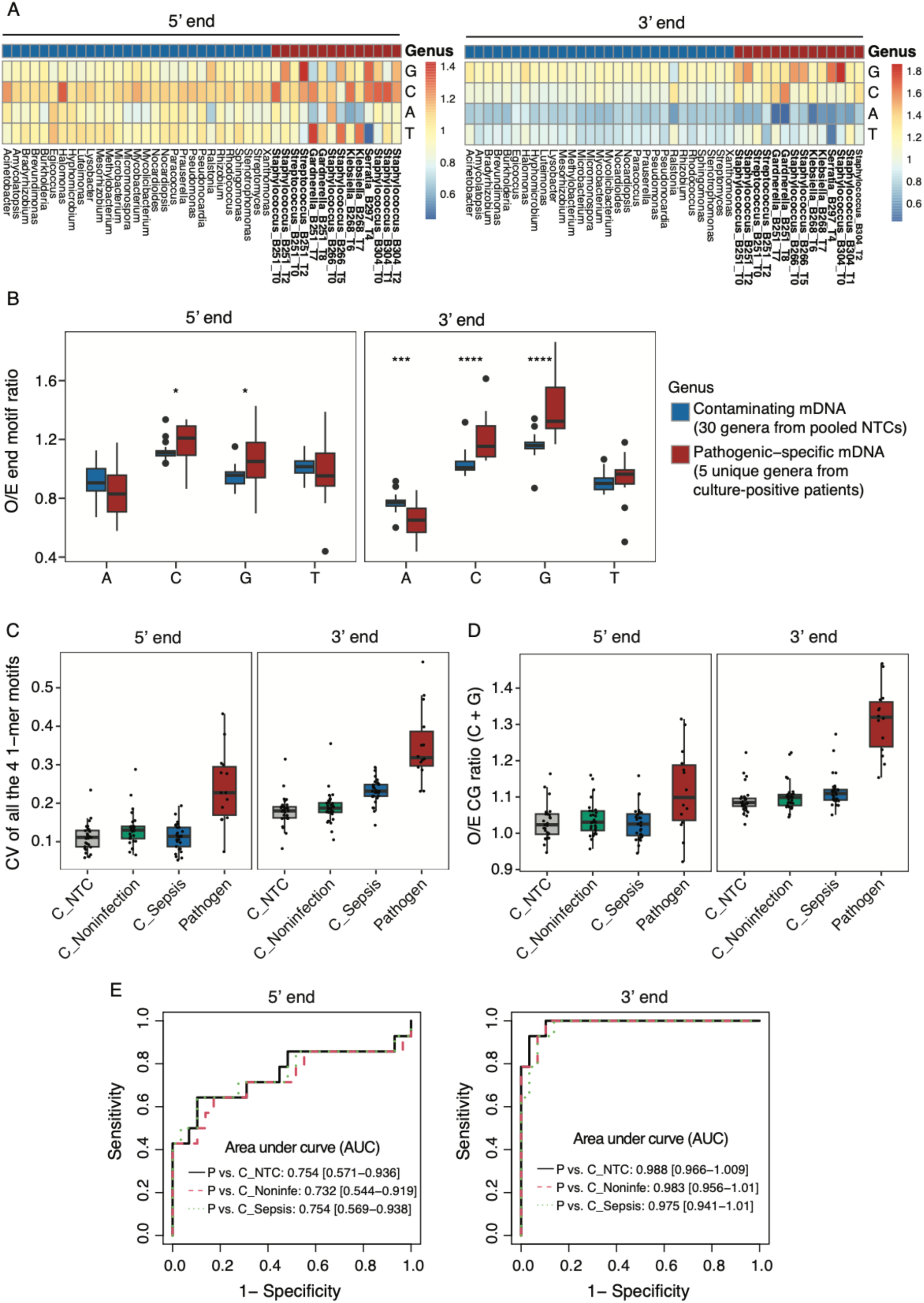
Mononucleotide (1-mer) microbial end motif patterns at the 5’ and 3’ fragment ends. (A) Heatmap shows the O/E ratios of 1-mer end motifs for contaminating genera and pathogenic genera. (B) Boxplots compare the O/E ratios of all 4 1-mer end motifs between pathogens and contaminants at both the 5’ and 3’ ends. (C) Coefficient of variation (CV) of 1-mer end motifs in culture-confirmed pathogens from sepsis samples and contaminants across pooled NTC, non-infection and sepsis groups. (D) The comparation of O/E CG ratios (combining C and G motifs) between pathogens and contaminants across pooled NTC, non-infection and sepsis samples. (E) ROC curves showing the performance of combined CG ratio in differentiating pathogens from contaminants across different groups. Contaminants are identified as the top30 most abundant genera from pooled NTCs. C_NTC, contaminants from pooled NTC samples; C_Noninfection, contaminants from pooled non-infection samples; C_Sepsis, contaminants from pooled sepsis samples.

The combined C and G end O/E ratio (CG ratio; Methods) remained stable and consistent across contaminants in NTC, non-infection and sepsis samples (Figure 5D), effectively distinguishing pathogen-derived mDNA from contaminants across all sample groups. Predictive modeling using the 1-mer based CG O/E ratio achieved AUCs of 0.75 and 0.98 for the 5’ and 3’ end, respectively (Figure 5E). Although the discriminatory performance of 1-mer analysis was slightly lower than that of the 2-mer approach, it still effectively differentiated pathogens from contaminants based on 3’ end features. The preservation of strong discriminatory power when transitioning from 2-mer to 1-mer end motif signatures demonstrates that the potential contaminants can be filtered out using 1-mer analysis in a reliable and cost-effective way.

## Discussion

A microbial DNA (mDNA) based sequencing assay has the potential to develop into a clinical test in the Intensive Care Unit for rapid diagnosis of sepsis once we overcome its technical limitations^13,15^. In an earlier study, we had found that combining ssDNA library preparation with size selection enriched for mDNA while depleting human cfDNA. While this approach increased the number of pathogen DNA fragments in the library, it also had a similar effect on DNA fragments from background contaminants, making it challenging to distinguish pathogen signal from background noise. In the current study, we tried to address this challenge and improve signal-to-noise ratio using analysis of end motifs in mDNA fragments. We observed that pathogen-derived mDNA is biased in the nucleotide motifs observed at both the 5’ and 3’ fragment ends compared with contaminants, with a stronger difference observed at the 3’ end. These differences were observed for both single and dinucleotide motifs, and enabled classification between common contaminants and culture-proven pathogens with more than 98% accuracy.

In the current study, we performed deep sequencing (∼120 million reads) on size-selected ssDNA libraries. Using a comparison of mDNA abundance alone between patients with infections and those without infections, sensitivity for pathogen detection was 38.5%. Interestingly, this method is sufficient for certain genera identified in microbiology cultures in this study. For example, *Gardnerella* and *Klebsiella* can be distinguished in sepsis samples from both non-infection samples and NTCs based on abundance alone. However, for other pathogens, specifically *Staphylococcus*, differentiation from commensals in non-infection samples remained challenging (Figure 2). The abundance of *Staphylococcus* cfDNA in sepsis samples overlapped substantially with that in non-infection samples, resulting in low detection rates when using abundance as the sole criterion. In such cases, analysis of microbial cfDNA fragmentation features could help pathogen-specific detection, especially for genera that would otherwise be false negative by abundance-based analysis.

Using 2-mer end motif analysis, we observed over-representation of CC and GG motifs at the 3’ end in culture-confirmed pathogens. Wang et al. ^19^ previously demonstrated that mDNA from infection-causing pathogens showed a preference for CC motif at the 5’ end, likely reflecting enzymatic degradation (e.g., DNASE1L3) of microbial cfDNA in plasma. In our study, although CC motif was enriched at the 5’ end of pathogen-derived microbial cfDNA compared to contaminants, this enrichment was not significant, potentially due to differences in anatomic site of infections and pathogen types between the two cohorts. Most culture-positive pathogens by Wang et al were from blood cultures and extra-pulmonary sites (e.g. liver and biliary), with *Klebsiella* and *Escherichia* being the most prevalent pathogens. In contrast, all infections in our cohort originated from respiratory sources, with positive cultures obtained from bronchoalveolar lavage (BAL), tracheal aspirate or sputum. *Staphylococcus* was the most common pathogen in our ICU patients and not present in the previous study. It is hypothesized that cfDNA end profiles are associated with specific nuclease activities and their sequence cleavage preferences^29–31^. Whether differences in enzymatic degradation across infection sites contribute to the difference of observed end motif patterns remains unclear. In addition, we also found a greater difference in motif patterns between pathogens and contaminants at the 3’ end than at the 5’ end. Since most library preparation methods modify or remove native 3’ ends, information about nuclease cleavage preference at the 3’ end remains limited. Further studies investigating the nuclease activity and cleavage patterns at both fragment ends will be important to better understand these processes.

Given low microbial biomass in our cohort (median reads per million, RPM: 0.12-37.3), we used 1-mer end motif analysis to reduce the number of fragments required for accurate motif characterization. The discriminatory performance of the 1-mer based analysis was slightly lower than that of the 2-mer based approach but remained effective in differentiating pathogens from contaminants (AUC = 0.98 for the 3’ end using the 1-mer approach). Notably, pathogen burden in our cohort was lower than reported in earlier studies (RPMs: 1.3-1189.4 in Wang et al.; 6-12010 in He et al.)^19,32^, and therefore, ssDNA and size selection for mDNA enrichment was particularly well suited. In samples with higher microbial abundance, less sequencing depth would be required to obtain sufficient reads for end motif analysis. Based on these findings, one possible clinical application would be to apply shallow metagenomic sequencing for initial screening. In cases where pathogens commonly present as contaminants (e.g., *Staphylococcus*) are detected, follow-up deep metagenomic sequencing combined with end motif analysis could aid clinical interpretation.

Although our findings can inform further development of microbial end motif analysis in size-selected ssDNA libraries for plasma metagenomic studies, several limitations should be noted. First, our sample size was limited to 11 critically-ill trauma patients enrolled prospectively. Larger cohorts are needed to determine whether the observed microbial end motif patterns can be generalizable across different infection and different pathogen types (including viral and fungal infections) and across infections at different anatomic sites. Second, although 3’ end features can better differentiate (compared to 5’) pathogen-derived mDNA from contaminating DNA, distinguishing true infection from colonization remains challenging. Future work will expand the current analysis to compare DNA fragment end motifs from candidate pathogen genera between no-template controls, infection free patients, healthy individuals and patients infections to determine if differences in end-motifs can potentially be utilized to improve pathogen detection as well as direct identification (without prior reliance on microbial cultures).

## Conclusions

We demonstrated that fragment end motif patterns in enriched single-stranded plasma microbial cfDNA are biased in pathogens compared to contaminants at both 5’ and 3’ ends. Fragmentation features of microbial cfDNA from size-selected ssDNA libraries provide new insights into 3’-end cleavage patterns and may aid in distinguishing pathogens from contaminants. This approach could enable the clinical implementation of plasma mDNA metagenomic sequencing as an effective biomarker for pathogen detection and identification.

## Abbreviations

cfDNA: cell-free DNA
mDNA: microbial cell-free DNA
dsDNA: double-stranded DNA
ssDNA: single-stranded DNA
ICU: intensive care unit
NTC: no-template control.

## Acknowledgments

This study was supported by American Association for the Surgery of Trauma (to M.T. Kisat), NIH National Institute of General Medical Sciences Award K08GM148858 (to M.T. Kisat), and the Wisconsin Partnership Program’s Postdoctoral Grant (to H. Zhang). We acknowledge the Department of Surgery Office of Clinical Research for their support in patient enrollment and sample processing.

## Competing interests

Eddie G. Dominguez and Mehreen T. Kisat are inventors on a non-provisional patent application submitted through the Wisconsin Alumni Research Foundation at the University of Wisconsin, Madison for “Methods for Enriching Microbial Cell-Free DNA in Plasma.” All other authors declare that they have no competing interests.

## Data availability

Genomic sequencing data reported here will be deposited in dbGaP upon acceptance of manuscript. Code reproducing the reported analysis will be deposited to Zenodo upon acceptance of manuscript.

## Supplementary figure legends

**Figure S1. Total microbial fraction across sepsis, non-infection, and no-template control (NTC) samples.** Boxplots show the total microbial fraction in 20 sepsis samples, 18 non-infection samples and 6 NTC samples. The total microbial fraction was calculated as the ratio of fragments classified at the phylum level to the total number of filtered sequencing fragments.

**Figure S2. Differential end motifs (DEMs) between pathogen-derived and contaminant-derived cfDNA**. Volcano plots show fold change (x-axis) and p-value (y-axis) for all the 16 dinucleotide end motifs comparing culture-confirmed pathogens from sepsis with contaminants from pooled NTCs at the 5’ and 3’ fragment ends. Red dots indicate motifs with significantly higher O/E ratios in pathogens, blue dots indicate motifs with significantly lower O/E ratios and gray dots represent not-significant differences.

**Figure S3. CG O/E ratios of pathogen-derived and contaminant-derived cfDNA across all samples.** Boxplots show CG O/E ratios for pathogens from sepsis samples and contaminants (top 30 genera in pooled NTCs) across all analyzed samples. Results are shown separately for the 5’ and 3’ end.

## Notes

### Author Declarations

Ethics committee/IRB of the University of Wisconsin-Madison gave ethical approval for this work

