## Supplemental Table and Figure for "Fragment end motif analysis to distinguish pathogens from contaminants in enriched plasma microbial DNA"

Table S1: List of all microbiology culture results for first 10 days of hospital admission.

| Patient | Hospital Day | Sample | Culture Site | Microbiology Culture Results |
| --- | --- | --- | --- | --- |
| B251 | 2 | T0 | Urine | No growth |
| B251 | 3 | None | Blood | No growth |
| B251 | 3 | None | Blood | No growth |
| B251 | 3 | None | Tracheal Aspirate | Moderate *Staphylococcus aureus*  Moderate *Streptococcus pneumoniae* |
| B251 | 4 | T2 | Urine | No growth |
| B251 | 9 | T7 | Blood | No growth |
| B251 | 9 | T7 | Blood | No growth |
| B251 | 9 | T7 | BAL | No growth |
| B251 | 9 | T7 | Urine | No growth |
| B251 | 10 | T8 | BAL | 5 x 10^3 CFU/mL *Staphylococcus aureus*  1 x 10^6 CFU/mL *Gardnerella vaginalis* |
| B266 | 2 | T0 | BAL | 4 x 10^5 CFU/ml *Staphylococcus aureus* |
| B266 | 2 | T0 | Blood | No growth |
| B266 | 2 | T0 | Blood | No growth |
| B266 | 6 | T4 | BAL | No growth at 10^3 CFU/mL. |
| B268 | 1 | T0 | Urine | No growth |
| B268 | 5 | T4 | Blood | No growth |
| B268 | 5 | T4 | Blood | No growth |
| B268 | 6 | T5 | BAL | 1 x 10^5 CFU/mL *Klebsiella (Enterobacter) aerogenes* |
| B268 | 6 | T5 | Urine | No growth |
| B268 | 10 | T9 | Blood | No growth |
| B268 | 10 | T9 | Blood | No growth |
| B297 | 6 | T5 | Urine | No growth |
| B297 | 6 | T5 | Blood | No growth |
| B297 | 6 | T5 | Blood | No growth |
| B297 | 6 | T5 | Sputum | Many *Haemophilus influenza*, few *Serratia marcescens/ureilytica* |
| B297 | 7 | T6 | BAL | 1 x 10^3 CFU/mL *Haemophilus influenzae* |
| B297 | 7 | T6 | Blood | No growth |
| B297 | 8 | T7 | Urine | No growth |
| B304 | 2 | T0 | Urine | No growth |
| B304 | 2 | T0 | Blood | No growth |
| B304 | 2 | T0 | Blood | No growth |
| B304 | 2 | T0 | Sputum | Moderate to many *Staphylococcus* aureus |
| B304 | 3 | T1 | BAL | 6 x 10^3 CFU/mL *Staphylococcus aureus* |

Figure S1

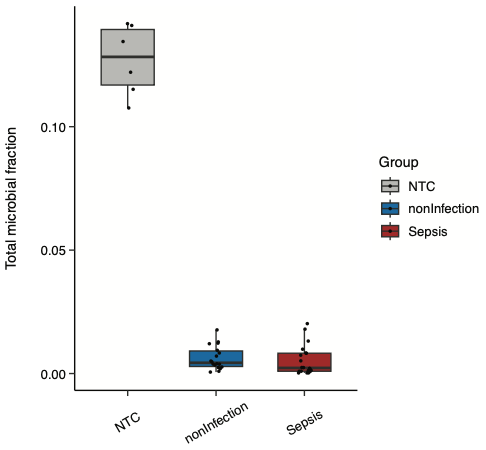

Figure S2

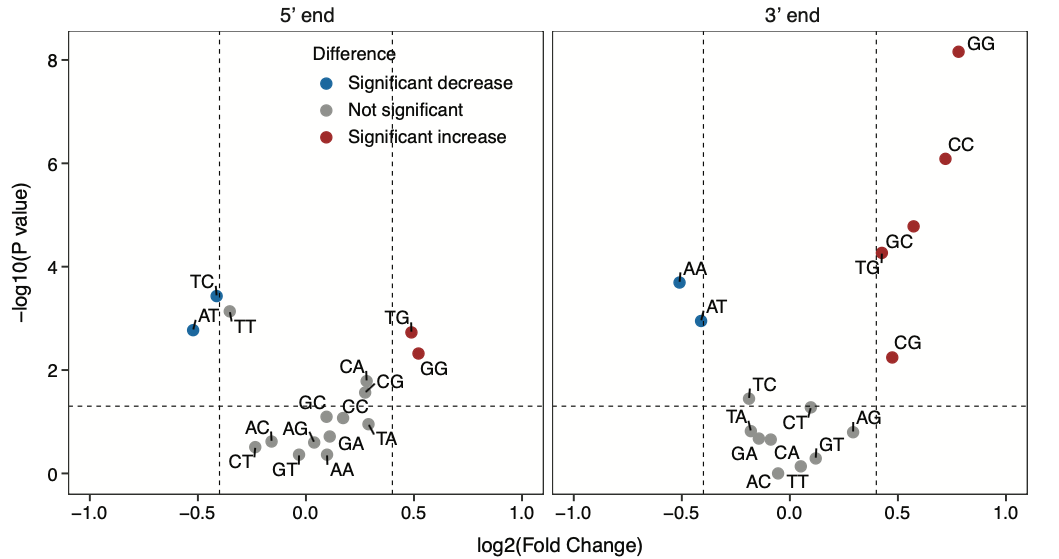

Figure S3

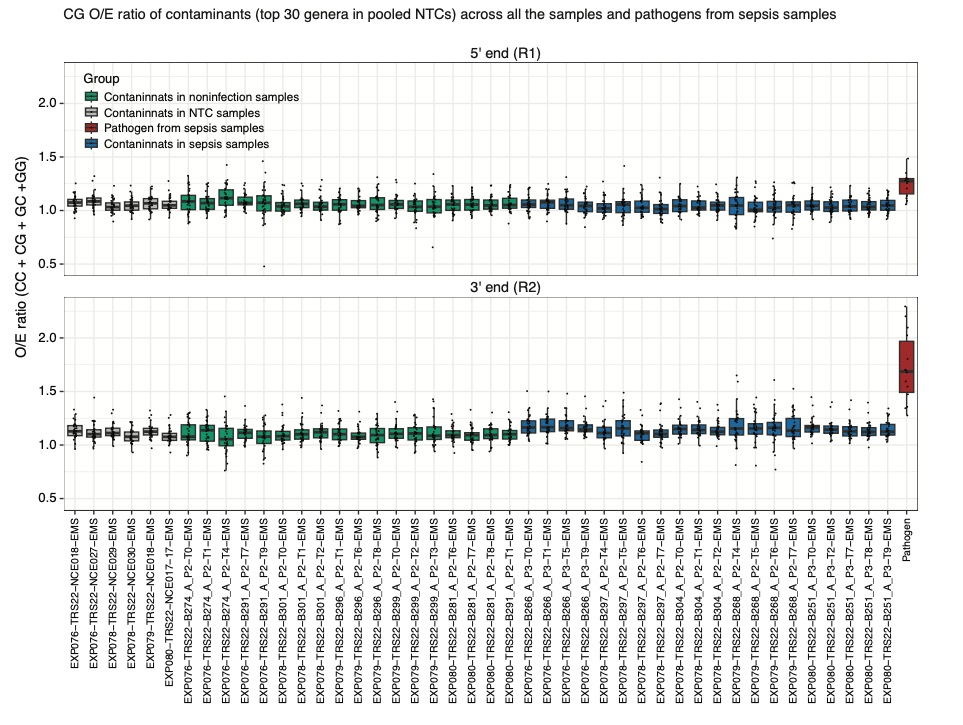
